# Genomic epidemiology reveals the dominance of Hennepin County in transmission of SARS-CoV-2 in Minnesota from 2020-2022

**DOI:** 10.1101/2022.07.24.22277978

**Authors:** Matthew Scotch, Kimberly Lauer, Eric D. Wieben, Yesesri Cherukuri, Julie M Cunningham, Eric W Klee, Jonathan J. Harrington, Julie S Lau, Samantha J McDonough, Mark Mutawe, John C. O’Horo, Chad E. Rentmeester, Nicole R Schlicher, Valerie T White, Susan K Schneider, Peter T Vedell, Xiong Wang, Joseph D Yao, Bobbi S Pritt, Andrew P Norgan

## Abstract

SARS-CoV-2 has had an unprecedented impact on human health and highlights the need for genomic epidemiology studies to increase our understanding of virus evolution and spread, and to inform policy decisions. We sequenced viral genomes from over 22,000 patient samples tested at Mayo Clinic Laboratories between 2020-2022 and use Bayesian phylodynamics to describe county and regional spread in Minnesota.

The earliest introduction into Minnesota was to Hennepin County from a domestic source around January 22, 2020; six weeks before the first confirmed case in the state. This led to the virus spreading to Northern Minnesota, and eventually, the rest of the state. International introductions were most abundant in Hennepin (home to the Minneapolis/St. Paul International (MSP) airport) totaling 45 (out of 107) over the two-year period. Southern Minnesota counties were most common for domestic introductions with 19 (out of 64), potentially driven by bordering states such as Iowa and Wisconsin as well as Illinois which is nearby. Hennepin also was, by far, the most dominant source of in-state transmissions to other Minnesota locations (n=772) over the two-year period.

We also analyzed the diversity of the location source of SARS-CoV-2 viruses in each county and noted the timing of state-wide policies as well as trends in clinical cases. Neither the number of clinical cases or major policy decisions, such as the end of the lockdown period in 2020 or the end of all restrictions in 2021, appeared to have impact on virus diversity across each individual county.

**Importance:** We analyzed over 22,000 SARS-CoV-2 genomes of patient samples tested at Mayo Clinic Laboratories during a two-year period in the COVID-19 pandemic that included Alpha, Delta, and Omicron VoCs to examine the roles and relationships of Minnesota virus transmission.

We found that Hennepin County, the most populous county, drove the transmission of SARS-CoV-2 viruses in the state after including the formation of earlier clades including 20A, 20C, and 20G, as well as variants of concern Alpha and Delta. We also found that Hennepin County was the source for most of the county-to-county introductions after its initial introduction with the virus in early 2020 from an international source, while other counties acted as transmission “sinks”. In addition, major policies such as the end of the lockdown period in 2020 or the end of all restrictions in 2021, did not appear to have an impact on virus diversity across individual counties.

## INTRODUCTION

Genomic epidemiology has provided valuable insight into transmission, evolution, and public health surveillance of severe acute respiratory syndrome coronavirus 2 (SARS-CoV-2), the cause of coronavirus disease 2019 (COVID-19). This has been feasible, in large part, to unprecedented viral genomic sequencing efforts across the globe. As of the 25^th^ of April 2023, there are over 15.4M virus sequences in GISAID [1] and over 6.8M in NCBI Virus [2] and GenBank [3]. Studies that focus on localized spread such as counties or regions within a state or province can highlight and uncover transmission events that could inform statewide surveillance and prevention efforts. However, there have been limited SARS-CoV-2 genomic epidemiology studies at this geographic level in the United States. Work by Moreno *et al*. [4] examined evolution and spread of SARS-CoV-2 among two counties (Dane and Milwaukee) in Wisconsin, from the start of the pandemic until the end of April 2020, based on analysis of 247 new full-length SARS-CoV-2 genomes combined with sequences in GISAID. Using this data, they derived county data on synonymous and non-synonymous single nucleotide variants (SNVs), and performed a variety of phylogenetic analyses (using Nextstrain [5] and BEAST2 [6]), determined R0 for their region, and examined the number and timing of introductions to the two counties and how each introduction subsequently impacted the local transmission [4]. The authors were ultimately able to conclude that early transmission within Dane County was not due to its initial introduction followed by local spread, but rather multiple later introductions into the region [4]. In other work, Deng *et al*. [7] sequenced 36 clinical samples from different counties in Northern California, sourced from the California Department of Public Health, Santa Clara County Public Health Department, and the University of California San Francisco. Their phylogenetic analysis revealed multiple California clusters including Santa Clara County, Solano County, San Benito County, as well as lineages from Washington State and Europe [7]. They also identified several early notable SNVs including D614G in the Spike protein [7]. Work by Müller *et al*. [8] examined SARS-CoV-2 introduction and spread in the State of Washington including at the county level early in the pandemic from February to July 2020 with a focus on the 614G variant and Google workplace mobility data. Alpert *et al*. created county-level risk maps for international importation of early Alpha variants via air transport [9]. More recently, Smith *et al*. [10] examined transmission of the Omicron variant among four counties in Arizona. Other work such as Valesano *et al*. [11], Holland *et al*. [12], Currie *et al*. [13], and Srinivasa et al. [14] examined spread on college campuses.

Although these studies have provided within-state snapshots into the genomic epidemiology of SARS-CoV-2, they did not consider how localized evolution and spread within a state changed over multiple years of the pandemic with the introduction and circulation of different variants of concern such as Alpha, Delta, and Omicron. Here, we leveraged amplicon-based high-throughput sequencing (HTS) and Bayesian phylodynamics to analyze the evolution and spread of SARS-CoV-2 into, and within, the State of Minnesota to understand the roles of specific counties and regions in transmission of the virus over a two-year period and across different viral clades, variants of concern (VoC), and state-wide mandates and policies.

## RESULTS

We sequenced SARS-CoV-2 genomes from genomic material collected from clinical samples of patients tested for SARS-CoV-2 infection at Mayo Clinic Laboratories (Figs. S1-S2) over a two-year period from March 2020 to March 2022. We combined these sequences with additional genomes generated for surveillance purposes by the Minnesota Department of Health (MDH) and performed Bayesian phylodynamics to understand in-state spread as well as the impact and timing of introductions into the State of Minnesota (see Methods).

Most of the patients from whom we collected a biological specimen and generated a SARS-CoV-2 genome resided in the State of Minnesota (96%) (Table S1). The breakdown by gender was nearly 50/50 between males and females while 50 percent of the patients were between 18-45. Fifteen percent were under 18 while 11 percent were 65 or older.

### Hennepin County consistently drives in-state transmission

We down-sampled Minnesota genomes nearly proportional to the number of COVID-19 cases per county and then added additional genomes from NCBI GenBank [3] as part of an international dataset used in Nextstrain [5] (see Methods). To address the computational burden of adding sequences to our already large dataset, we aggregated the additional samples into discrete traits *International* and *USA* and grouped counties with less sequences into areas in the state such as Southern, Central, and Northern Minnesota (Fig. 1 and Table S2).

**Figure 1.**
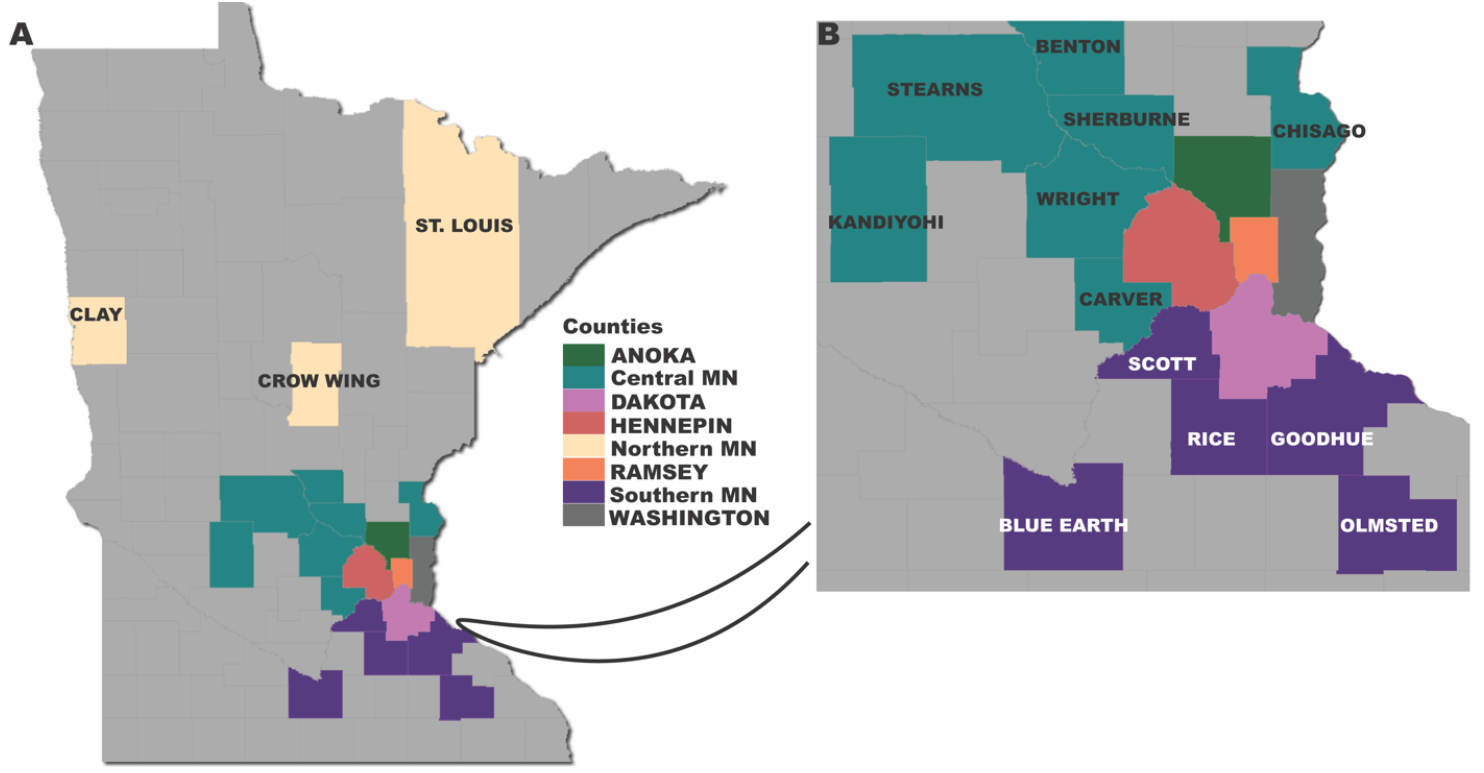
Map of Minnesota counties included in the phylodynamic analysis. Counties with the same color are part of a discrete region (northern, central, and southern) used in the analysis. We show an inset of a portion of the map for visualization purposes.

We implemented Bayesian phylodynamic models to examine transmissions in Minnesota from early 2020 to early 2022 (see Methods). We recorded Markov jumps [15] to estimate the timing of introductions and their directionality. After introductions from domestic and international locations, our analysis shows that Hennepin County, the most populous county which includes Minneapolis, the most populated city, drove the transmission of SARS-CoV-2 viruses in the state (Fig. 2). This includes the formation of earlier clades including 20C and 20G, as well as variants of concern Alpha and Delta (Fig. 2A). The counties in *Central Minnesota* contributed to spread including 20C, Alpha, and Delta, while *Southern Minnesota* contributed mostly to 20G.

**Fig. 2.**
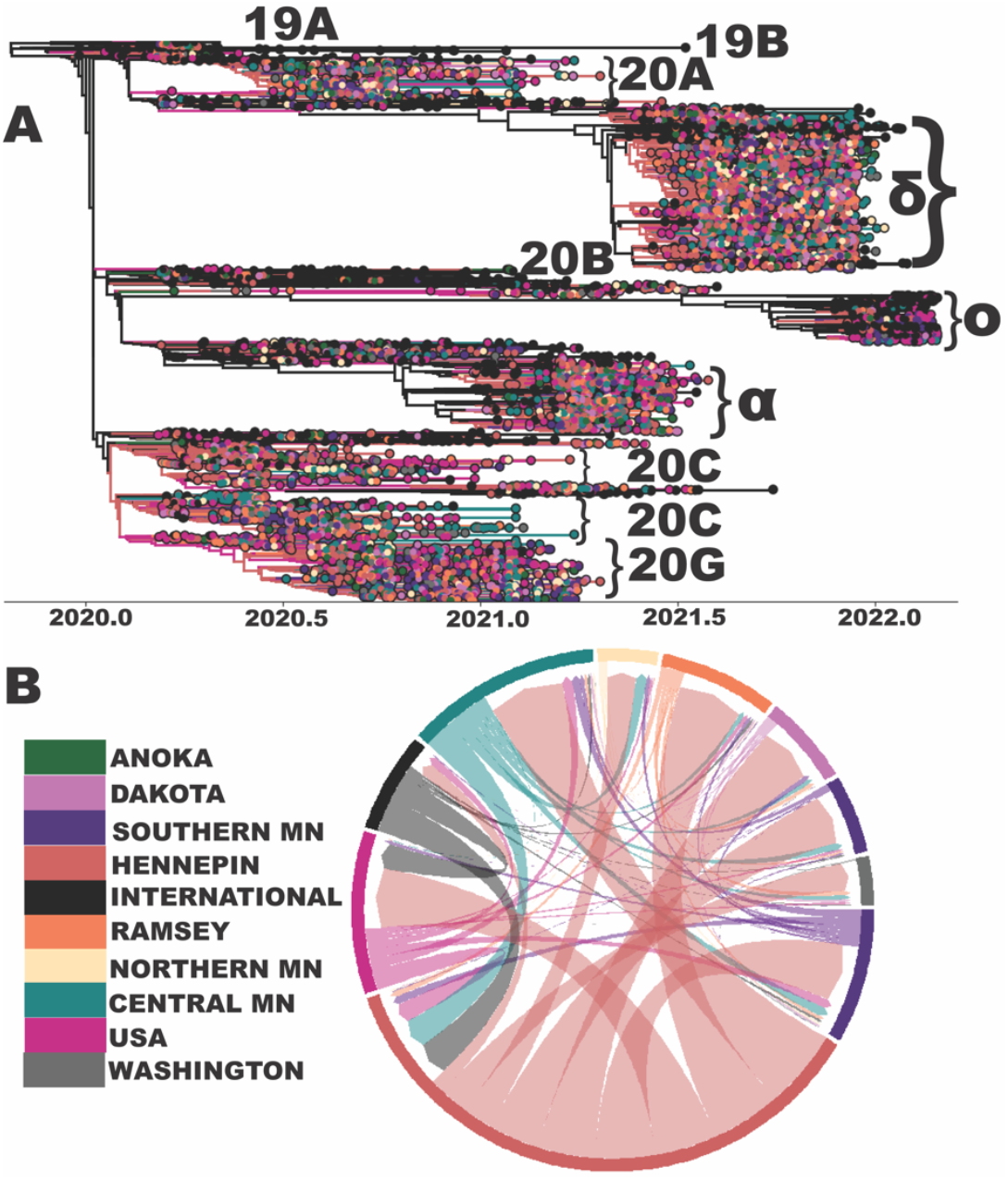
SARS-CoV-2 evolution and spread to and within the State of Minnesota. A) Maximum clade credibility (MCC) tree of 6,188 SARS-CoV-2 genomes from Minnesota counties and regions as well as international locations and other domestic locations in USA. We manually annotate clades by Nextclade-assigned names or VoCs [16] using their lowercase Greek letter. We do not label less represented VoCs in our tree such as Gamma or Epsilon. Some clades such as 20A are not monophyletic in the tree but we label their most populous clade. B) Markov jumps between locations as shown via a Chord diagram. The colors for both panels represent the locations depicted in the legend. *Central MN* includes seven Minnesota counties: Benton, Carver, Chisago, Kandiyohi, Sherburne, Stearns, and Wright. *Northern MN* includes three counties: Clay, Crow Wing, and Saint Louis. *Southern MN* includes five counties: Blue Earth, Goodhue, Olmsted, Rice, and Scott. *USA* includes all states except for Minnesota. Abbreviation: MN – Minnesota, α - alpha, δ - delta, ο - omicron. We combined panel A and B and re-created the legend and text labels in Adobe Illustrator for visualization purposes.

Markov jump estimates (Fig. 2B) as shown via a Chord diagram suggest that transmission of SARS-CoV-2 within the state largely originated from Hennepin County (thick arcs and wider fragments at the outer circle). However, we also note existence of transmission back to these areas (white space between arc points and outer fragment) from nearby counties from Central Minnesota.

We measured the ratio of introductions to total viral flow into and out of each county by month from March 2020 to January 2022. A value of *1* suggests a county as solely being a “sink” (accepts SARS-CoV-2 lineages but never exports them to other counties), while a value of *0* indicates a county as solely being a “source”. Anoka, Dakota, Ramsey, Northern Minnesota, Southern Minnesota, and Washington were fueled by introductions mostly throughout the pandemic (Fig. 3). Meanwhile, Central Minnesota (outside of Hennepin and Ramsey) was dominated by introductions early in the pandemic but later in 2020 experienced brief trends of higher virus exportation. Hennepin County showed a drastically different trend than all others as it consistently acted as a source for other Minnesota counties over the nearly two-year period. However, it did experience brief periods of fluctuation such as a spike in the ratio of introductions towards the end of 2020 and early 2021, potentially driven by the dominance of out of state introductions.

**Fig. 3.**
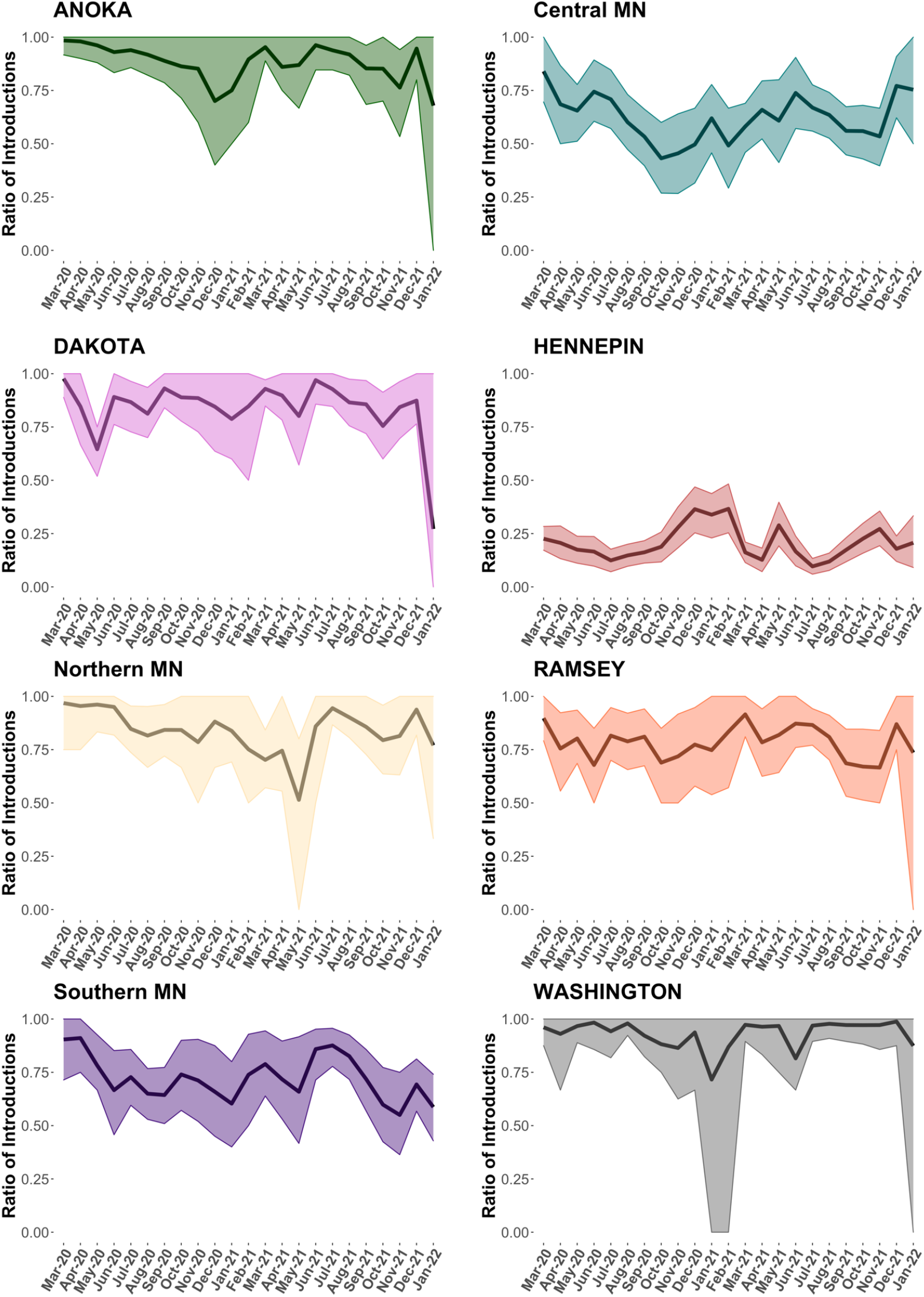
Ratio of introductions to total viral flow into and out of each discrete location by month from March 2020 – January 2022. We show the posterior mean ratio and 95% Bayesian highest posterior density interval. We note that some locations such as Anoka, Dakota, Ramsey, and Washington have wider intervals during certain months, such as January 2022, due to a decrease in local sampling.

### Low-to-intermediate spatial mixing within the State of Minnesota

We assessed county-specific virus diversity via a normalized Shannon diversity index (Fig. S3) that we computed based on the duration of time associated with continuous partitions of the phylogeographic tree as determined by Markov jumps [17] (see Methods). The index, in this context, measures the degree of spatial structure (based on counties) during the evolution and spread of SARS-CoV-2 viruses in Minnesota. A value of *0*, indicates exclusive spatial structure such as an outbreak contained to only one county [17]. Conversely, a value of *1*, suggests significant spatial mixing of SARS-CoV-2 between counties [17]. The counties and regions show low to intermediate (0.25 to 0.5 Shannon) spatial mixing with brief periods of waxing and waning. The two dotted vertical lines indicate changes in state-wide policy. The first vertical line indicates the end of lockdown in Minnesota on May 18, 2020 [18]. The second line on May 28, 2021 indicates the end of all COVID-19 restrictions in the state [19]. Neither of these policy decisions appeared to have significant impact on virus diversity across each individual county. Anecdotally, (looking at the trends of each graph) the changes in case counts over time does not appear to have a relationship with county-specific diversity.

### Hennepin County received the vast majority of out of state introductions and was the dominant source for in-state transmission

We focused on the timing and source of introductions into the state during the pandemic (Fig. 4) as estimated from our maximum clade credibility tree (Fig. 2A). The earliest introduction into Minnesota was to Hennepin County from a international source on around January 22, 2020 (depicted with an arrow in Fig. 4). This is about one month before the first patient in the state, a man from Ramsey County (which borders Hennepin), developed symptoms and around six weeks before (March 6, 2020) the Department of Health confirmed the infection [20]. The first county-to-county introductions were estimated to originate from Hennepin to somewhere in Northern Minnesota around February 22 and from Hennepin to Washington County (also in the northern part of the state) around February 24. International introductions were most abundant in Hennepin (home to the Minneapolis/St. Paul International (MSP) airport) totaling 45 (out of 107) over the two-year period. Southern Minnesota counties were most common for domestic introductions with 19 (out of 64), potentially driven by bordering states such as Iowa and Wisconsin as well as Illinois which is nearby. Hennepin also was, by far, the most dominant source of in-state transmissions to other Minnesota locations (n=772) over the two-year period.

**Fig. 4.**
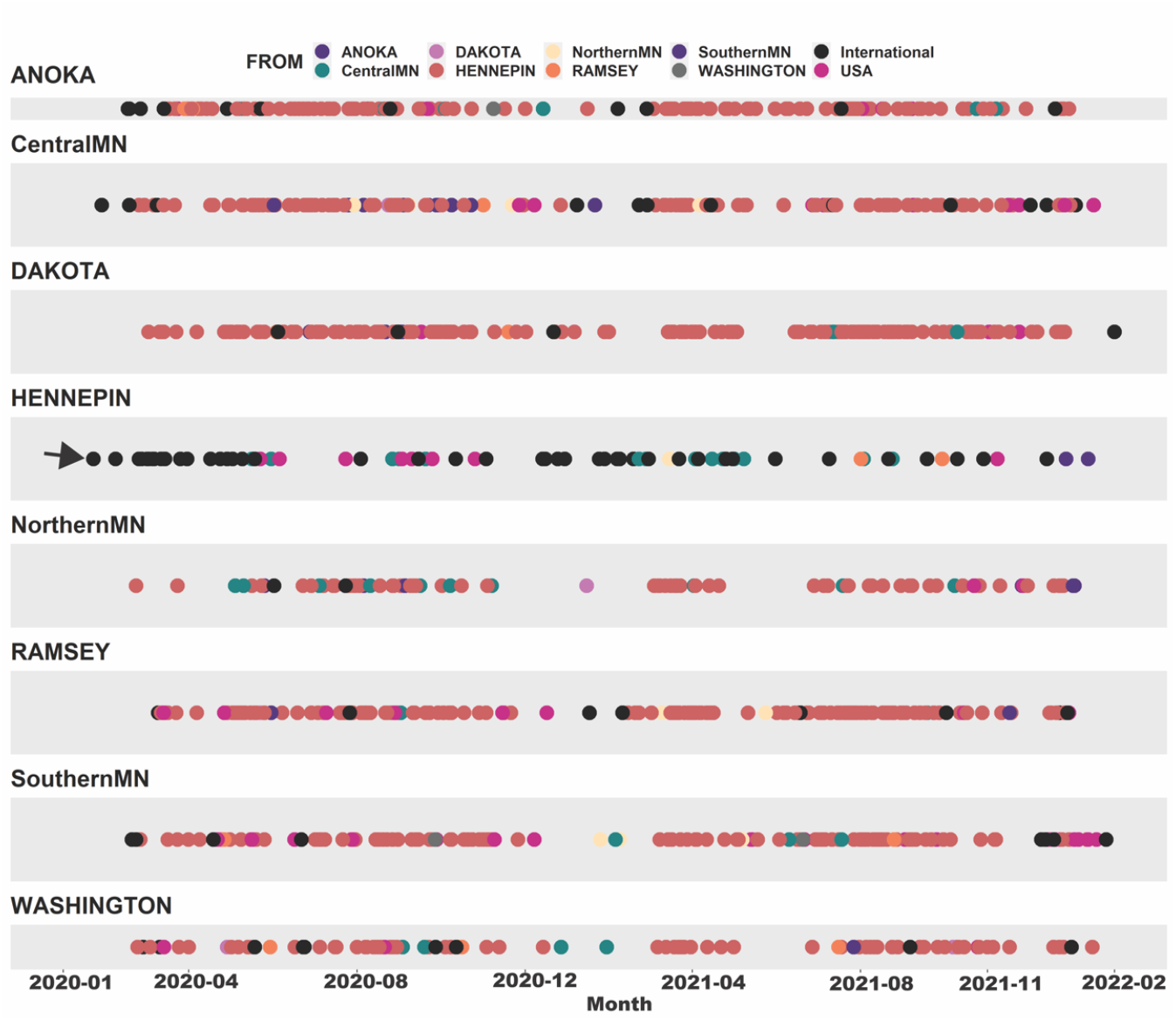
Timing and source of international, domestic, and within state introductions for each discrete location. The colors correspond to the source location. We use an arrow to show the first introduction into Minnesota which occurred in Hennepin County at around January 22, 2020 (from an international location). We used Baltic to extract introductions (migration events) along the annotated branches of the phylogeographic tree for node states with a posterior probability of ≥ 0.90. We combined the location panels into one figure in R and replaced the original month labels (x-axis) in Adobe Illustrator for visualization purposes.

## DISCUSSION

We analyzed over 22,000 new genomes of patients tested at Mayo Clinic Laboratories during a two-year period in the COVID-19 pandemic. We focused our analysis on in-state transmission of SARS-CoV-2, mostly at the county (2^nd^ administrative boundary) level, to describe the spread into and within Minnesota. Despite numerous efforts in genomic epidemiology, few studies have focused on county-to-county transmission in the US over most of the pandemic (including different VoCs). We expand on earlier efforts such Moreno *et al*. and [4] and Deng *et al*.[7] but include multiple variants and an extensive timeframe. We found that spread in the state was dominated by viruses from Hennepin County, which contains the largest metropolis, and that other regions including northern and southern Minnesota acted mainly as “sinks” for in-state transmission.

The earliest introduction into Minnesota was to Hennepin from an international source about six weeks before the first confirmed case in the state. This suggests that earlier (and likely milder) infections of SARS-CoV-2 occurred before the first documented case. Interestingly, while Hennepin drove in-state transmission, it did not result in variations of location-specific spatial diversity. We found that all counties and regions had low-to-intermediate (0.25 to 0.5 Shannon) spatial mixing with brief periods of waxing and waning. The fluctuation in spatial diversity over time (that did exist) did not appear to be impacted by key state-mandated policies nor did it appear to have any relationship with reported clinical cases (Fig. S3).

As the virus continues to evolve, more within state genomic epidemiology studies are needed to inform local and state public health response by highlighting the roles of various counties on state-wide transmission. In addition, they can elucidate the impact of out of state introductions on local spread which can inform policies such as travel.

We note several limitations in the study including the likelihood of location-specific sampling bias. We attempted to supplement known locations of patients in our study (biased towards southeastern Minnesota) with existing sequences provided via the Minnesota Department of Health. We initially scaled our number of sequences to the rate of known COVID-19 cases, and, after doing so, omitted counties with a limited number of sequences (as well as outliers of sequences from included counties). Thus, we are unable to account for virus spread from less populated areas of the state. We also attempted to include a representative sample of USA and international sequences. However, it is possible that additional sequences (context) might change the distribution of virus clades and the timing of introductions into the state, which could alter our interpretations of SARS-CoV-2 spread. We also only included early Omicron sequences and thus we are unable to describe an informed picture of its evolutionary diffusion in the state. In addition, our use of different versions of the DRAGEN pipeline over the course of our two-year study period, likely led to differences in variant frequencies across virus lineages/VoCs.

## MATERIALS and METHODS

### RNA extraction, library preparation and next generation sequencing

From March 2020 to March 2022, we analyzed patient nasopharyngeal or mid-nasal turbinate swabs that tested positive for COVID-19 via RT-qPCR at Mayo Clinic Laboratories and had a Ct value of 28 or lower. We extracted viral RNA on the Hamilton Microlab STAR Automated Liquid Handler system (Hamilton Company, Reno, NV USA) with the use of Promega Maxwell HT Viral TNA Kit (Fitchburg, WI). We generated libraries using the COVIDSeq Test reagent kit from Illumina (San Diego, CA USA) following the manufacturer’s instructions. We sequenced the pooled libraries as 100 × 2 paired end reads using the NovaSeq SP sequencing kit and Xp 2-Lane kit with NovaSeq Control Software v1.6.0. We used the Illumina RTA version 3.4.4 for base-calling.

We de-multiplexed raw sequence data into individual sample fastq files using bcl2fastq2-v2.19.0 [21]. We used Illumina’s Dynamic Read Analysis for GENomics (DRAGEN) COVID Lineage software and pipeline [22] (versions 3.5.1,3.5.3, and 3.5.6) for reference-based alignment to Wuhan-1 (NC_045512.2), quality assessment, variant calling, and generation of consensus sequences.

We excluded sequences from downstream analysis if they met any of the following criteria, including: 1) given an overall score of *fail* by the DRAGEN pipeline due to having an insufficient amount of detectable viral reads; 2) given an overall quality score by Nextclade [16] as *bad;* 3) potentially contaminated based on presence of unusual allele frequencies (< 0.9); 3) duplicate runs; 4) positive or negative controls.

### Phylogenetic analysis of SARS-CoV-2

We assembled a representative dataset (n = 6,188; Fig. S4) that included SARS-CoV-2 genome sequences from the 20 counties with the greatest number of reported COVID-19 cases as of February 28, 2022 as well as a global representation of sequences available via GenBank as part of an open access dataset from Nextstrain (Table S2) [23]. We used the list of accessions to download sequences from NCBI Virus [2]. We removed sequences less than 29K nucleotides in length as well as duplicates.

We included sequences from December 2019 (including Wuhan-1, GenBank accession MN908947) to February 28, 2022, as well as their sampling location and collection date metadata. To partially address sampling bias, we sampled at a rate of 5 sequences per 1,000 county cases and used the *filter* module in augur [24] to distribute (as equally as possible) our heterochronous sequences by month (Fig. S4). For each county, we attempted to include SARS-CoV-2 genomes across the two-year timeframe by supplementing our dataset with sequences provided by the Minnesota Department of Health (MDH) (and available in GISAID). The MDH sequences were produced from randomly selected samples from clinics and community testing sites. Sample Ct values were equal or below 30.

We aligned all sequences using Mafft [25] and used trimAl v1.4.rev22 [26] to remove columns which contained more than 70% gaps. We created initial an phylogenetic tree via Nextstrain’s *augur tree* command [5] with IQTree and rooted the tree based on Wuhan-1 (MN908947) [27]. We used TempEST [28] to examine the temporal signal of our heterochronous samples and removed one sequence as an outlier. We used *augur refine* and the *keep-root* option to modify our tree with sequence metadata. We removed additional sequences that had potential misassigned clades or produced inconsistences with the phylogenetic structure as shown in Nextstrain’s global all-time subsampled dataset [29].

### Phylodynamics of SARS-CoV-2 in Minnesota

We used R package *ape* [30] to confirm that our starting tree was rooted and non-bifurcating in order to comply with our downstream inferencing framework. For Bayesian inference, we leveraged a pre-release of BEAST v1.10.5 (ThorneyTreeLikelihood v0.1.1) and BEASTGen v0.3 (pre-thorney) to specify a more efficient likelihood function intended for larger sequence datasets [31, 32]. We used our starting tree and a non-parametric Bayesian SkyGrid coalescent model for our tree prior [33]. We ran two Markov-chain Monte Carlo (MCMC) simulations each for 5 × 10^8^ steps and sampling every 5 × 10^4^ steps. We combined these two runs via LogCombiner v.10.4 [34] after removing 10% burn-in. We checked for convergence of model parameters via Tracer v1.7.1 [35] with an ideal effective sample size (ESS) threshold of 200. We generated log marginal likelihoods and evaluated population growth priors via a stepping stone and path sampling procedure [36]. Our results suggested the use of the non-parametric Skygrid tree prior over a constant growth model (Table S3).

We used LogCombiner to sample 1,000 trees from the posterior distribution and used this as empirical data for ancestral state reconstruction of our location traits. We specified all non-US sequences as “International” and non-Minnesota US states as “USA”. For computational efficiency, we kept the five counties with the greatest number of cases as independent locations and grouped the remaining fifteen counties into three discrete regions including Southern, Central, and Northern Minnesota (Table S2). In BEAUti [34], we specified an asymmetric transmission rate matrix of *K(K*1)* where *K* is equivalent to the number of discrete locations (n = 10 for our dataset). We recorded Markov jumps [15] between locations to estimate the timing and source of introductions and specified a MCMC of 5 × 10^6^ sampling every 5 × 10^2^ steps. We used TreeAnnotator v.10.4 [34] to create a single maximum clade credibility (MCC) tree after 10% burn-in. We used baltic [37] for tree visualization and to extract the timing of discrete location transitions along the branches of the MCC for our estimates of introductions. We excluded transmission chains with low support of discrete origin and destination names by including only nodes with a posterior probability of ≥ 0.90 for the location state. We used SpreadD3 [38] to calculate the Bayes factors to identify the most parsimonious origin-destination scenarios (Table S4).

We used two programs of the BEAST library [39], introduced in [17], as part of our Bayesian phylodynamic analyses. *TreeMarkovJumpHistoryAnalyzer* samples from the posterior distribution of trees to collect the timing and location of each Markov jump [17]. We used the output from this program to calculate the ratio of introductions to total viral flow into and out of each county (number of introductions / (number of introductions + number of exports)) as described in Lemey *et al*. [17] as well as the visualization of the weights of pairwise transmission between counties via a chord diagram. *TreeStateTimeSummarizer*, which also samples from the posterior distribution of trees, notes the contiguous partitions for a given discrete state [17]. We used the output from this program to calculate the normalized Shannon diversity metric as described in Lemey *et al*. [17]. We used this measure to assess the level of location diversity for the viruses within each county during a specified time-period. For our analysis, we used *NormShannon method in* the R package *QSutils* [40] to calculate normalized monthly diversity metrics for each county and *HDinterval [41]* for the corresponding 95% highest posterior density region.

### Human subjects and ethics approval

This research was conducted under approval of ethics by the Mayo Clinic Institutional Review Board and assigned a study ID IRB#: 20-005896 and entitled *Large Scale Whole Genome Sequencing of SARS-CoV-2*. All our published datasets contain randomly generated study IDs and no personal identifiers. All data analysis was performed behind the Mayo Clinic firewall.

## Data Availability

We have deposited the SARS-CoV-2 genomes and metadata from this study in GISAID with a list available at doi.org/10.55876/gis8.220720me. The Minnesota Department of Health sequences used in this study are available on GISAID with acknowledgments at doi.org/10.55876/gis8.220709mv. Our GenBank international sequences were identified via the Nextstrain site and obtained from NCBI Virus. We have deposited BEAST XML files, empirical set of posterior trees, and our introductions in figshare at 10.6084/m9.figshare.21777995; 10.6084/m9.figshare.21778004; 10.6084/m9.figshare.21777998; 10.6084/m9.figshare.22679449.

## Data availability

We have deposited the SARS-CoV-2 genomes and metadata from this study in GISAID with a list available at doi.org/10.55876/gis8.220720me. The Minnesota Department of Health sequences used in this study are available on GISAID with acknowledgments at doi.org/10.55876/gis8.220709mv. Our GenBank international sequences were identified via the Nextstrain [23] site and obtained from NCBI Virus [2]. We have deposited BEAST XML files, empirical set of posterior trees, and our introductions in figshare at <u>10.6084/m9.figshare.21777995</u>; <u>10.6084/m9.figshare.21778004; 10.6084/m9.figshare.21777998</u>; <u>10.6084/m9.figshare.22679449</u>.

## Acknowledgements

This publication was supported by funding from the Center for Individualized Medicine-Mayo Clinic Research. Research reported in this publication was also supported by the National Institute of Allergy and Infectious Diseases (NIAID) of the National Institutes of Health (NIH) under Award Number R01AI164481 (to MS). The content is solely the responsibility of the authors and does not necessarily represent the official views of the National Institutes of Health. The authors would like to thank Gytis Dudas for his assistance with the Baltic Python library. The authors would also like to thank Scott Lunt and Rob Timmer for their assistance with the mforge high-performance computing environment. The authors acknowledge the laboratories that submitted the SARS-CoV-2 genome data to GISAID that were used in this work.

The authors dedicate this publication to Mr. Peter T Vedell who sadly passed away during the writing of this manuscript.

## Author contributions

MS designed the genomic epidemiology study, performed the data analysis, and wrote the manuscript. APN and BSP jointly supervised the study and providing funding, reviewed the manuscript, and approved the final manuscript. CER performed data analysis, assisted in reviewing and writing the manuscript. KL, YC, PTV and EWK implemented the DRAGEN and Nextstrain bioinformatics pipeline and variant analysis pipelines, performed variant analysis, and reviewed the manuscript. JMC, JSL, MM, SJM, VTW, NRS, SKS and EDW supervised and performed the sample extraction, viral genomic sequencing, primary analysis of the sequencing and variant data, and reviewed the manuscript. JJH managed the execution of the study and reviewed the manuscript. JCO extracted the clinical data, performed primary data analysis, and reviewed the manuscript. JDY coordinated the provision of samples for quality control and sequencing, assisted with variant interpretation, and reviewed the manuscript.

XW supervised the generation of the Minnesota Department of Health genomic sequences, performed primary analysis of the MDH sequence data, and reviewed the manuscript.

## Materials & Correspondence

Please direct requests for materials or other correspondence to Andrew P. Norgan, MD, Mayo Clinic, 200 First Street SW, Rochester, MN USA 55905.

## SUPPLEMENTARY MATERIALS

**Fig. S1.**
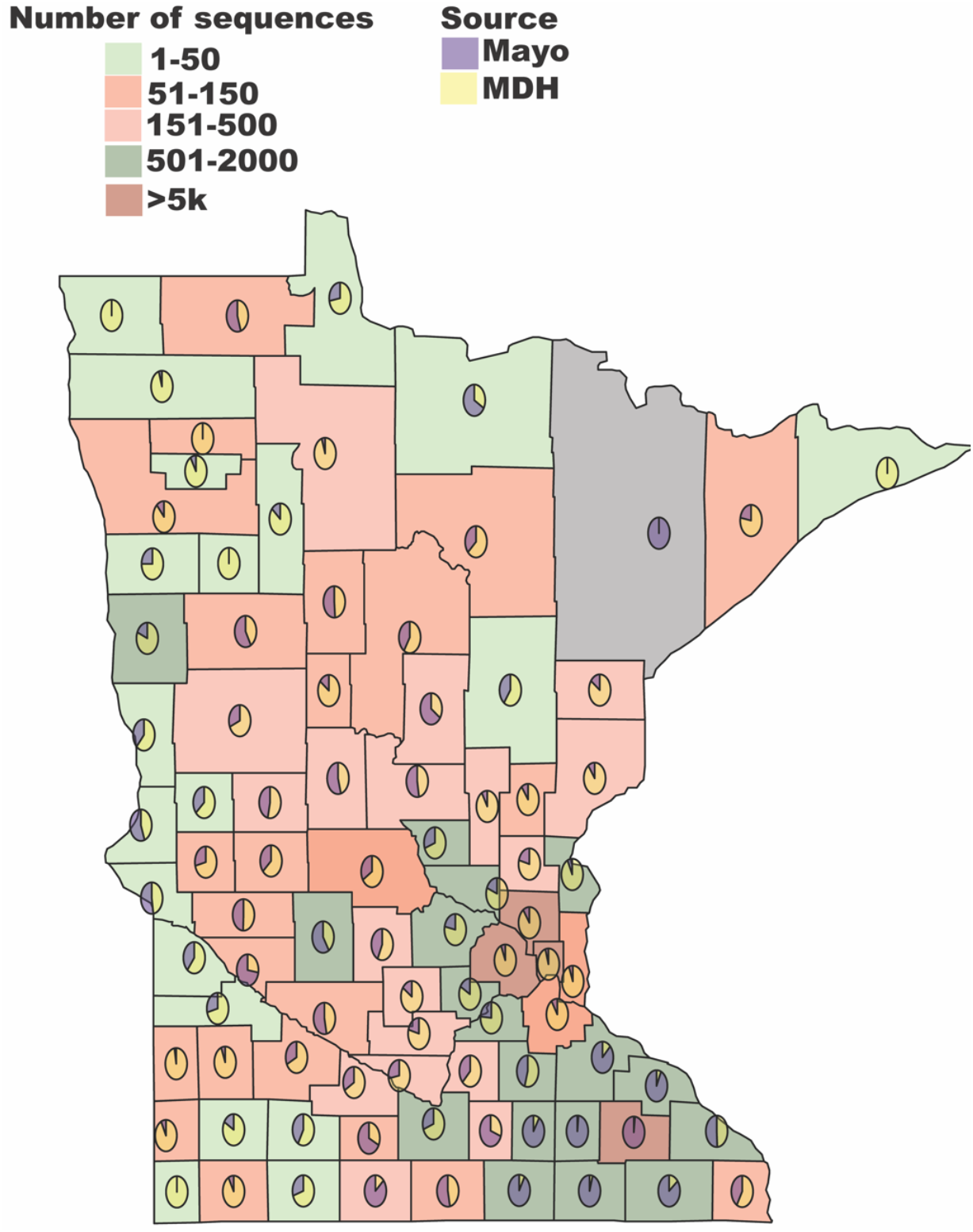
County map of Minnesota with number of sequences (N=76,875) eligible for analysis by source. Here, *Mayo* (N=21,669) represents new sequences generated from this study at Mayo Clinic Laboratories with a known sampling location in a Minnesota county. MDH (N=55,206), refers to a sequence available on GISAID with county metadata provided by the Minnesota Department of Health (MDH). We re-created the legend and text labels in Adobe Illustrator for visualization purposes.

**Fig. S2.**
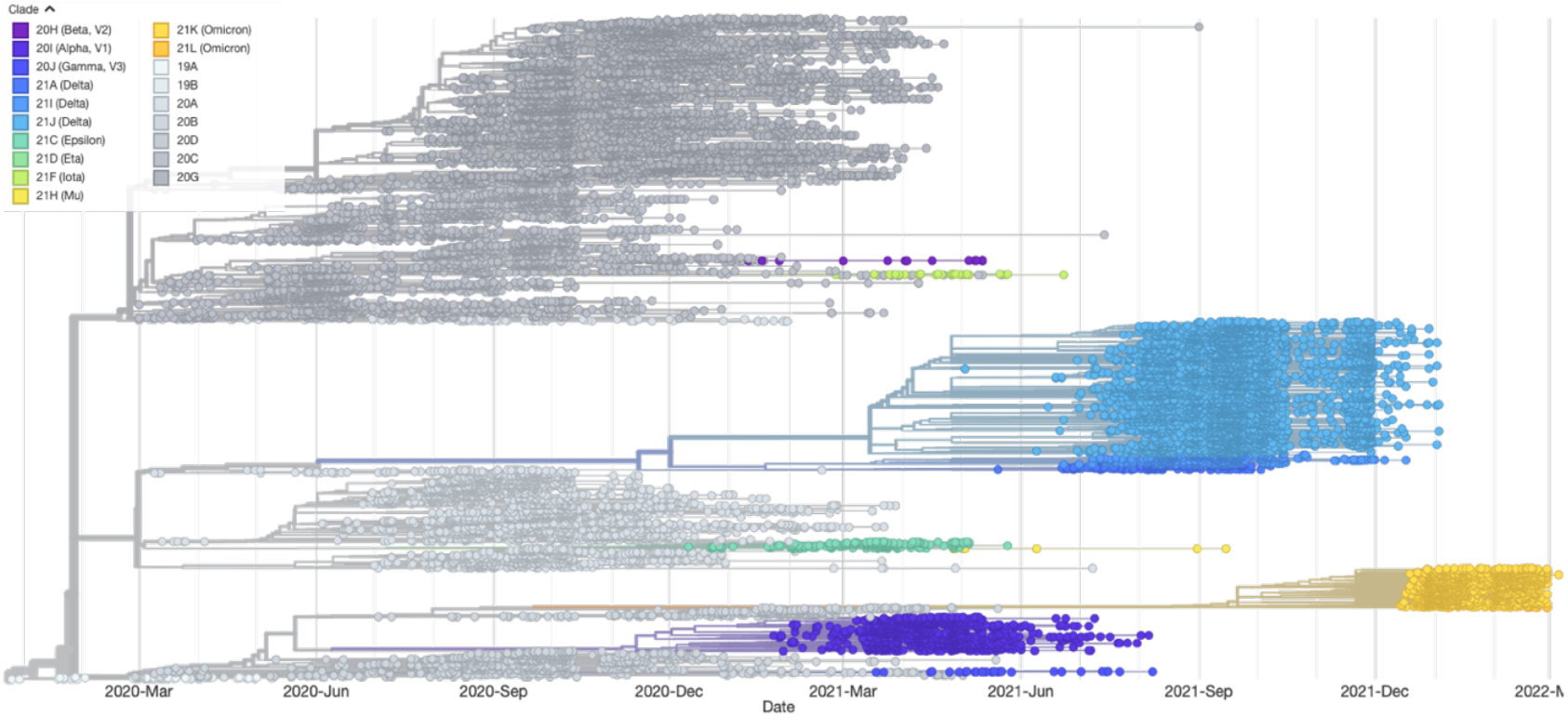
Phylogeny of 24,070 full genome SARS-CoV-2 sequences generated for this study from 2020-2022 via Nextstrain (augur v15.0.2). Branch colors indicate the assigned clade.

**Fig. S3.**
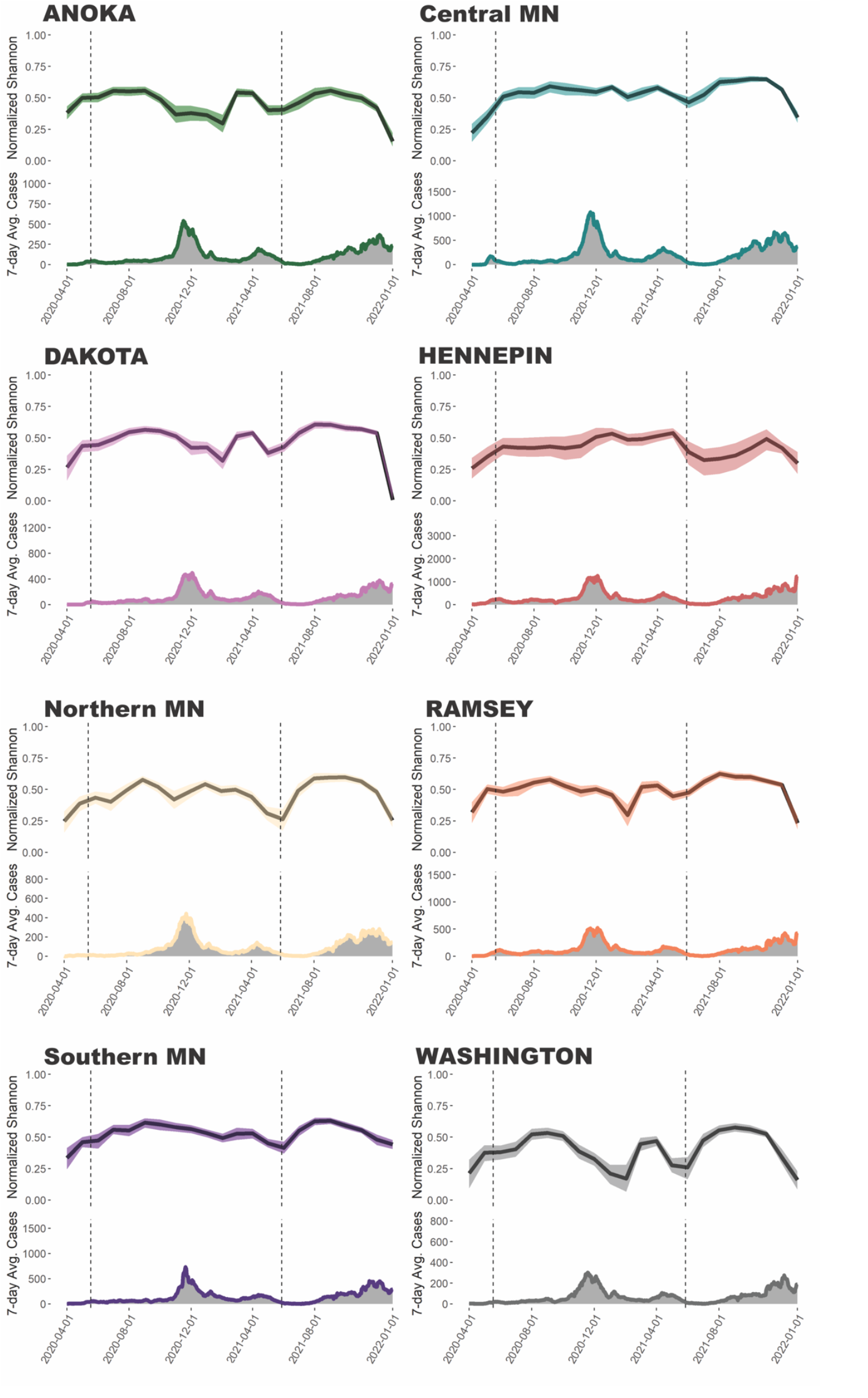
Virus diversity and cases per county/location. On the top of each panel, we show the normalized Shannon diversity index over time for each Minnesota county or region in the study. The shaded areas represent the 95% Bayesian highest posterior density (HPD). Below, we show the seven-day average cases for the particular county or region obtained from [42] via outbreak.info. We aggregated the data for Northern, Central, and Southern Minnesota based on the counties listed in Table S2. The first vertical line indicates the end of lockdown in Minnesota on May 18, 2020 [18]. The second vertical line indicates the end of all COVID-19 restrictions in the State on May 28, 2021 [19]. We combined location-specific graphs and re-created the location labels in Adobe Illustrator for visualization purposes.

**Fig. S4.**
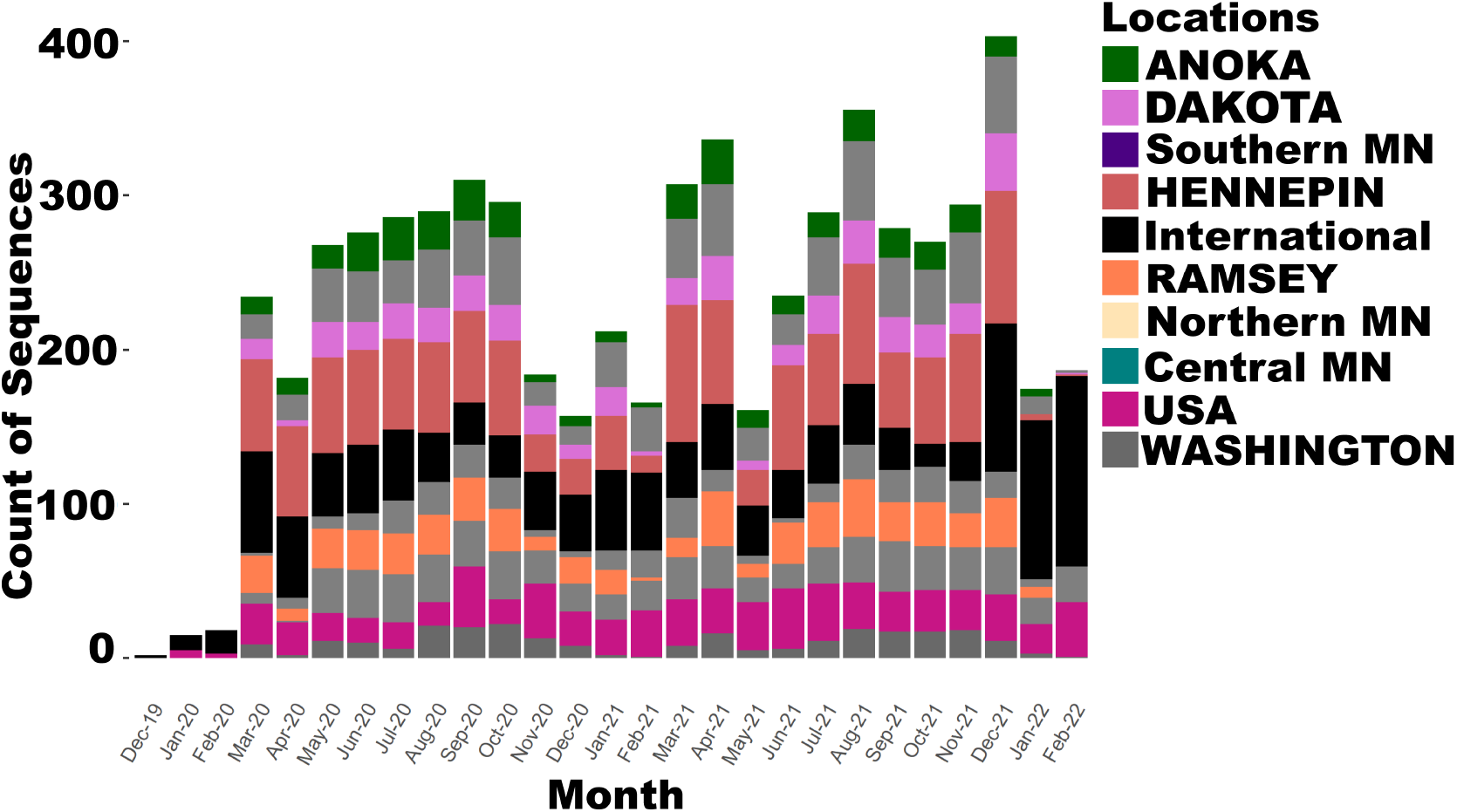
Sequence distribution (n = 6,188) by Minnesota county/region by month for our phylodynamic analysis. We re-created the legend and text labels in Adobe Illustrator for visualization purposes.

**Table S1.** Demographics of N = 22,514 Mayo Clinic Laboratories COVID-19 patients included in the study.

| **Demographic** | **Category** | **Count (%)** |
| --- | --- | --- |
| Gender | Male | 11,423 (49) |
|  | Female | 11,076 (51) |
|  | Non-binary | 15 (<1) |
| Age | < 18 | 3,416 (15) |
|  | 18-45 | 11,204 (50) |
|  | 46-64 | 5,350 (24) |
|  | 65+ | 2,544 (11) |
| State | Minnesota | 21,669 (96) |
|  | Wisconsin | 413 (2) |
|  | Iowa | 184 (1) |
|  | Other States | 148 (1) |

**Table S2.**
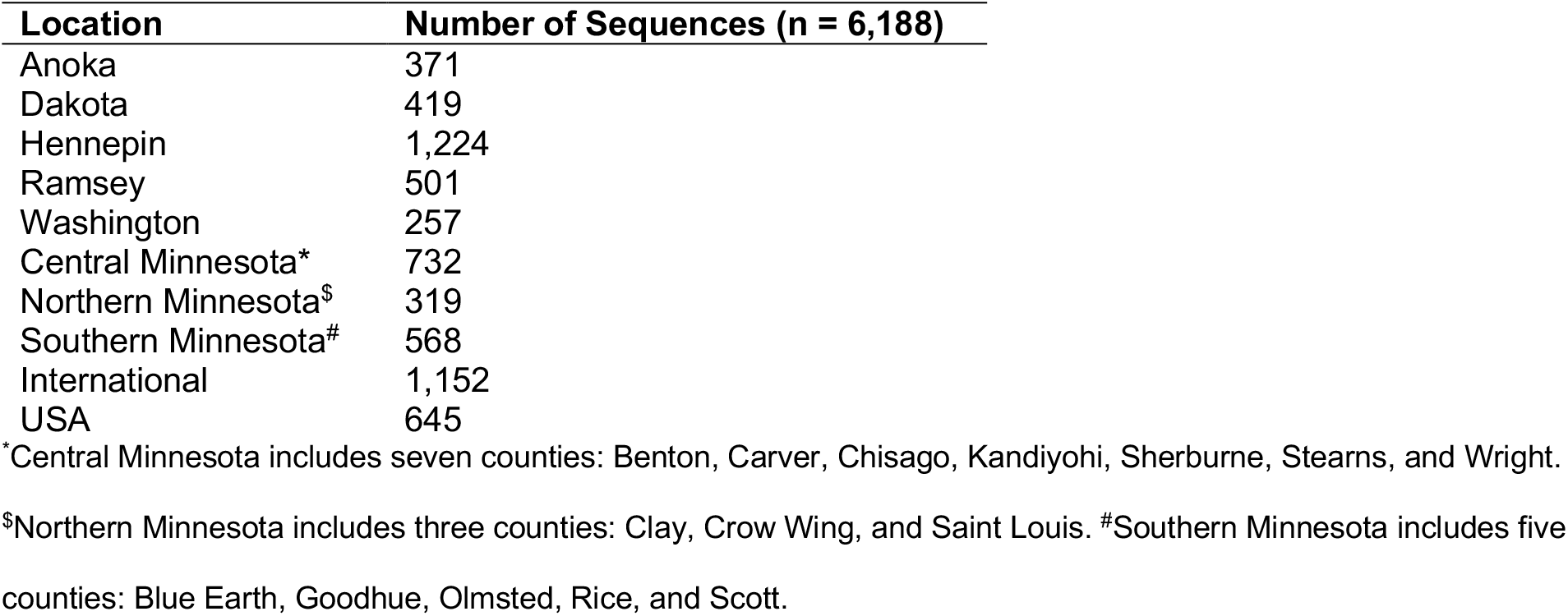
Number of sequences by location for the phylodynamic analysis.

**Table S3.** Log marginal likelihood values calculated via stepping-stone and path-sampling for different coalescent models under a strict molecular clock. The results favor the use of a non-parametric Skygrid coalescent model.

| **Tree Prior** | **Stepping-Stone** | **Path-Sampling** |
| --- | --- | --- |
| Skygrid | -12,690.78 | -12,683.44 |
| Constant | -13,041.41 | -13,033.6 |

**Table S4.** Bayes factor and posterior probability values for support of non-zero SARS-CoV-2 transmission of Minnesota counties that met our *a priori* threshold > 100. We highlight the 48 routes out of 90 in this category in grey. We omitted routes that did not involve Minnesota such as USA→International or International →USA. Hennepin was most often included as the origin (9/48). Meanwhile, Southern MN was most popular as the destination (8/48), followed by Ramsey (7/48) and Central MN (7/48).

| From | To | Bayes Factor | Posterior Probability |
| --- | --- | --- | --- |
| Central MN | HENNEPIN | 74,564.19 | 1 |
| Central MN | Northern MN | 74,564.19 | 1 |
| DAKOTA | RAMSEY | 74,564.19 | 1 |
| DAKOTA | WASHINGTON | 74,564.19 | 1 |
| HENNEPIN | International | 74,564.19 | 1 |
| HENNEPIN | Northern MN | 74,564.19 | 1 |
| HENNEPIN | RAMSEY | 74,564.19 | 1 |
| HENNEPIN | Southern MN | 74,564.19 | 1 |
| HENNEPIN | USA | 74,564.19 | 1 |
| HENNEPIN | WASHINGTON | 74,564.19 | 1 |
| International | USA | 74,564.19 | 1 |
| RAMSEY | WASHINGTON | 74,564.19 | 1 |
| Central MN | ANOKA | 74,564.19 | 1 |
| HENNEPIN | ANOKA | 74,564.19 | 1 |
| HENNEPIN | Central MN | 74,564.19 | 1 |
| Southern MN | Central MN | 74,564.19 | 1 |
| USA | Central MN | 74,564.19 | 1 |
| HENNEPIN | DAKOTA | 74,564.19 | 1 |
| International | HENNEPIN | 74,564.19 | 1 |
| Southern MN | HENNEPIN | 74,564.19 | 1 |
| USA | HENNEPIN | 74,564.19 | 1 |
| USA | International | 74,564.19 | 1 |
| Southern MN | Northern MN | 74,564.19 | 1 |
| USA | Southern MN | 74,564.19 | 1 |
| Central MN | WASHINGTON | 37,277.95 | 1 |
| International | RAMSEY | 37,277.95 | 1 |
| Southern MN | DAKOTA | 37,277.95 | 1 |
| Northern MN | Southern MN | 12,420.46 | 0.999 |
| Central MN | Southern MN | 7,448.96 | 0.999 |
| USA | RAMSEY | 6,771.03 | 0.999 |
| RAMSEY | DAKOTA | 4,134.63 | 0.998 |
| Northern MN | Central MN | 2,477.46 | 0.997 |
| Southern MN | RAMSEY | 2,251.49 | 0.996 |
| Central MN | RAMSEY | 2,063.17 | 0.996 |
| ANOKA | Central MN | 1,954.15 | 0.996 |
| Northern MN | RAMSEY | 1,214.21 | 0.993 |
| RAMSEY | Central MN | 999.45 | 0.992 |
| Central MN | DAKOTA | 460.72 | 0.982 |
| Southern MN | USA | 403.72 | 0.98 |
| USA | ANOKA | 320.23 | 0.975 |
| RAMSEY | ANOKA | 285.31 | 0.972 |
| RAMSEY | HENNEPIN | 276.34 | 0.971 |
| RAMSEY | Southern MN | 266.89 | 0.97 |
| DAKOTA | Central MN | 229.21 | 0.965 |
| DAKOTA | Southern MN | 213.66 | 0.963 |
| WASHINGTON | Southern MN | 187.44 | 0.958 |
| USA | WASHINGTON | 148.05 | 0.947 |
| International | Southern MN | 136.24 | 0.943 |
| International | Central MN | 95.72 | 0.92 |
| USA | Northern MN | 58.84 | 0.877 |
| Southern MN | International | 51.76 | 0.862 |
| Southern MN | ANOKA | 42.38 | 0.836 |
| DAKOTA | Northern MN | 32.64 | 0.798 |
| Northern MN | DAKOTA | 30.72 | 0.788 |
| International | ANOKA | 30.1 | 0.784 |
| International | WASHINGTON | 20.69 | 0.714 |
| ANOKA | RAMSEY | 16.33 | 0.663 |
| Northern MN | HENNEPIN | 14.37 | 0.634 |
| WASHINGTON | ANOKA | 12.64 | 0.604 |
| ANOKA | HENNEPIN | 7.95 | 0.49 |
| Southern MN | WASHINGTON | 3.65 | 0.306 |
| Central MN | USA | 3.48 | 0.296 |
| DAKOTA | HENNEPIN | 3.28 | 0.283 |
| ANOKA | DAKOTA | 2.47 | 0.23 |
| DAKOTA | ANOKA | 2.4 | 0.225 |
| ANOKA | WASHINGTON | 2.24 | 0.213 |
| RAMSEY | Northern MN | 2.13 | 0.205 |
| ANOKA | USA | 1.9 | 0.186 |
| ANOKA | Southern MN | 1.49 | 0.153 |
| Northern MN | USA | 1.47 | 0.151 |
| Northern MN | WASHINGTON | 1.3 | 0.135 |
| WASHINGTON | RAMSEY | 1.27 | 0.133 |
| WASHINGTON | DAKOTA | 1.17 | 0.123 |
| WASHINGTON | HENNEPIN | 1.15 | 0.121 |
| RAMSEY | USA | 1.1 | 0.117 |
| WASHINGTON | Central MN | 0.93 | 0.101 |
| DAKOTA | USA | 0.83 | 0.091 |
| WASHINGTON | Northern MN | 0.72 | 0.08 |
| International | DAKOTA | 0.68 | 0.076 |
| Northern MN | International | 0.64 | 0.072 |
| Central MN | International | 0.43 | 0.049 |
| USA | DAKOTA | 0.4 | 0.047 |
| RAMSEY | International | 0.28 | 0.033 |
| International | Northern MN | 0.28 | 0.032 |
| WASHINGTON | International | 0.25 | 0.03 |
| WASHINGTON | USA | 0.24 | 0.028 |
| ANOKA | Northern MN | 0.17 | 0.02 |
| DAKOTA | International | 0.15 | 0.017 |
| ANOKA | International | 0.13 | 0.016 |
| Northern MN | ANOKA | 0.11 | 0.013 |

